# Missed Appointments and Associations with Clinical Outcomes in A Large National Healthcare System

**DOI:** 10.64898/2026.03.28.26349531

**Authors:** Ying Yin, Yan Cheng, Youxuan Ling, Christopher Ruser, Hamada H. Altalib, Robin M. Masheb, Jeffrey Kravetz, Stuart J. Nelson, Ali Ahmed, Charles Faselis, Cynthia A. Brandt, Qing Zeng-Treitler

**Affiliations:** Veterans Affairs Medical Center, Washington, DC, USA; George Washington University, Washington, DC, USA; Georgetown University, Washington, DC, USA; VA Connecticut Healthcare System, West Haven, CT, USA; Yale School of Medicine, New Haven, CT, USA; Veterans Affairs Palo Alto Health Care System, Palo Alto, CA, USA; Stanford University School of Medicine, Palo Alto, CA, USA

## Abstract

**Importance:** Missed outpatient appointments, including no-shows and cancellations, may disrupt continuity of care and be associated with worse outcomes, but long-term system-wide patterns and clinical implications are not well characterized.

**Objective:** To characterize variation in missed appointment rates in the Veterans Health Administration (VHA) over time and by geographic location, visit modality, and preexisting conditions, and to evaluate associations between missed appointment rates and adverse outcomes among veterans with posttraumatic stress disorder (PTSD) or traumatic brain injury (TBI).

**Design:** Cohort study using VHA Corporate Data Warehouse outpatient appointment data from January 1, 2000, through December 31, 2024.

**Setting:** National integrated health care system of the VHA.

**Participants:** System analysis includes all scheduled outpatient appointments with a valid status, and outcome analysis includes veterans with PTSD (n = 1 429 890) or TBI (n = 554 553), diagnosed before 2023.

**Exposures:** For system -level analyses, missed appointment rates were calculated. In outcome analyses, 2023 missed appointment rates were categorized into tertiles within the cohort and appointment type.

**Main Outcomes and Measures:** One year risks of all-cause hospitalization, all-cause mortality, and hospitalization or death beginning January 1, 2024.

**Results:** Among 2,162,520,880 outpatient appointments from 2000 to 2024, 6.5% were no-shows and 25.4% were canceled. Across facilities, no-show rates ranged from 3.5% to 14.1%, patient-initiated cancellation rates from 9.7% to 26.0%, and clinic-initiated cancellation rates from 8.5% to 17.9%. In 2023, veterans with amputation, Parkinson disease, PTSD, or TBI had higher missed appointment rates than veterans without these conditions. Among veterans with PTSD, the highest no-show tertile, compared with none, was associated with higher mortality (HR, 1.91; 95% CI, 1.84-1.98) and hospitalization or death (HR, 1.07; 95% CI, 1.05-1.08). Among veterans with TBI, the highest no-show tertile was associated with hospitalization or death (HR, 1.65; 95% CI, 1.61-1.69).

**Conclusions and Relevance:** Missed outpatient appointments were common in the VHA and varied substantially across facilities and over time. Among veterans with PTSD or TBI, higher missed appointment rates, particularly no-shows, were associated with increased risks of hospitalization and mortality, suggesting that these patterns may help identify high-risk veterans for targeted outreach.

**Key Points:** *Question:* What patterns and sources of variation in missed outpatient appointments were observed in the Veterans Health Administration, and were higher missed appointment rates associated with one year hospitalization and mortality among veterans with PTSD or TBI?

*Findings:* In this cohort study of 2,162,520,880 outpatient appointments from 2000 to 2024, no-shows accounted for 6.5% and cancellations for 25.4%, with substantial temporal and geographic variation. Higher no-show rates were associated with increased hospitalization and mortality among veterans with PTSD or TBI.

*Meaning:* Higher missed appointment rates, particularly no-shows, may signal disengagement from care and elevated clinical risk, supporting targeted outreach for high-risk veterans.

## 1. Introduction

Missed patient appointments, including both no-shows and cancellations initiated by either clinics or patients, represent missed opportunities for care and pose a pervasive challenge in outpatient settings. Reported no-show rates vary widely, ranging from 5% to over 30%, depending on the specialty, location, and patient population.^1-3^ Within the Veterans Health Administration (VHA), a recent study reported that approximately 11% of primary care appointments were either canceled on the day of the visit or resulted in a no-show.^4^ While no-shows and cancellations are distinct phenomena, they both pose a challenge for healthcare providers, as they can disrupt clinic schedules, incur time and financial costs, and delay timely patient care or disease management.^5^

Prior research conducted largely outside the VA healthcare system has demonstrated that poor appointment adherence and treatment disengagement are associated with higher risks of relapse, hospitalization, and mortality.^6,7^ While not focused on U.S. Veterans, research in Traumatic Brain Injury (TBI) and Post Traumatic Stress Disorder (PTSD) shows that non-adherence to follow-up is associated with delayed recovery and greater symptom burden.^8,9^ The association between missed appointments and subsequent outcomes, however, has not been thoroughly examined across different clinical conditions, both within and outside the VA.

The VHA, as one of the largest integrated healthcare systems in the United States, provides a unique setting to examine missed appointment patterns at scale and to evaluate their clinical significance across diverse patient populations. In this study, we leverage more than two decades of VA outpatient appointment data to address three related aims. First, we characterize long-term temporal and geographic patterns of missed outpatient appointments across VA facilities. Second, we examine the association between selected veteran-prevalent conditions and missed appointment rates within a single year. Third, we evaluate whether higher missed appointment rates are associated with an increased one-year risk of adverse clinical outcomes, including hospitalization and mortality, with particular focus on post-traumatic stress disorder (PTSD) and traumatic brain injury (TBI), conditions that require ongoing outpatient follow-up.

Together, these aims are intended to provide a comprehensive view of missed appointments at both system and patient levels. We hypothesized that missed appointment rates would vary substantially across time and geography within the VA system and would be higher among Veterans with high-need pre-existing conditions. We further hypothesized that Veterans with higher missed appointment rates would experience worse subsequent clinical outcomes, consistent with the effects of disrupted continuity of care.

## 2. Methods

### 2.1 Overall Missed Appointment Patterns

#### 2.1.1 Data Source

To examine overall missed appointment rates and their geographic and temporal patterns across VA facilities, we included all outpatient appointments, both face-to-face and telehealth, scheduled between January 1, 2000, and December 31, 2024, using data from the VA Corporate Data Warehouse (CDW). A total of 2,265,982,879 outpatient appointments were initially identified. Appointments scheduled during inpatient stays and those marked as “no action taken” or with unclear status (N = 103,461,999) were excluded. The final analytic dataset included 2,162,520,880 outpatient appointments (**Online Figure 1A**).

**Figure 1.**
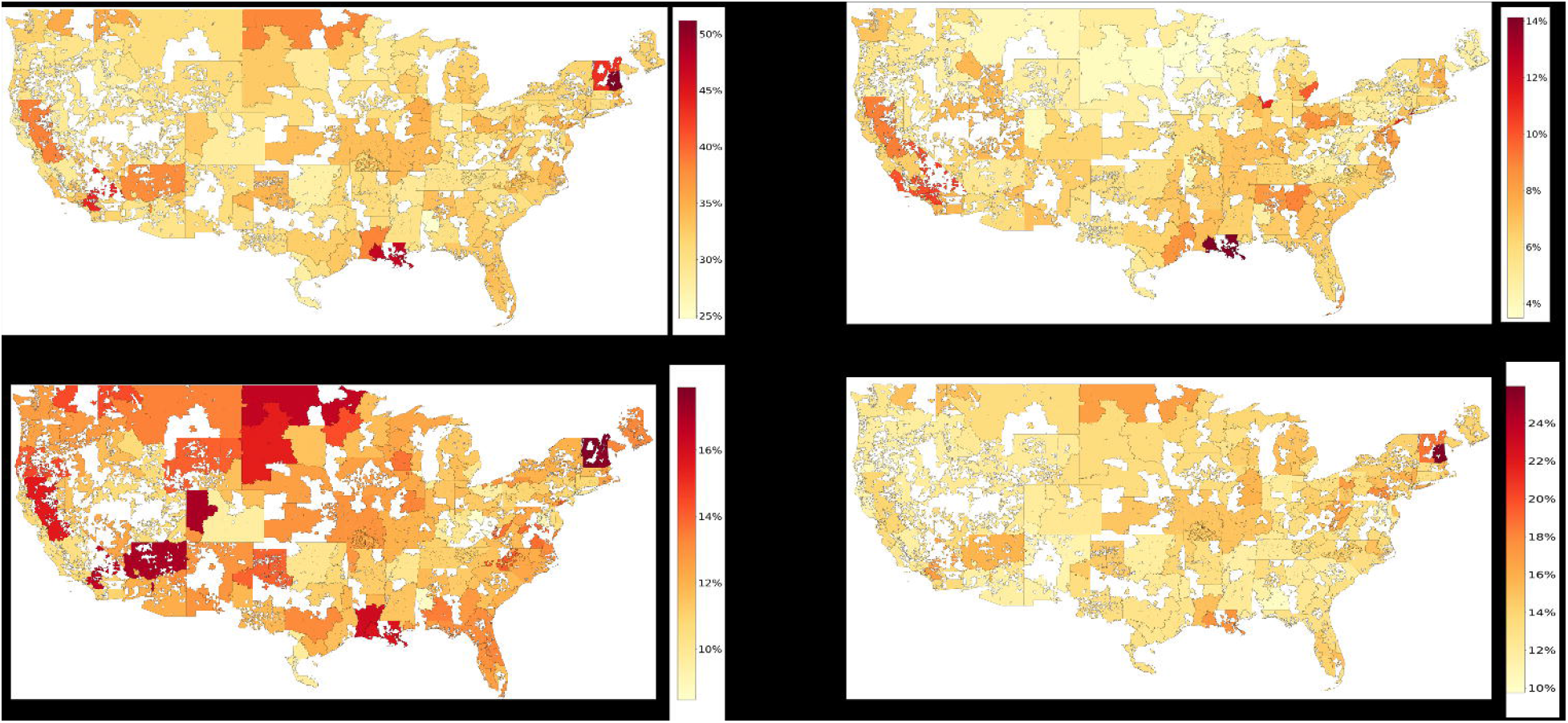
Percentage of all missed (A), no-show (B), clinic-initiated cancellation (C), and patient-initiated cancellation (D) appointments, 2000–2024, by VA station.

#### 2.1.2 Missed Appointment Measures

Missed appointment outcomes were categorized into three distinct types: no-shows, clinic-initiated cancellations, and patient-initiated cancellations. To assess trends, appointment-level rates were calculated longitudinally across study years or aggregated by VA facility at the Station Number (STA3N) level.

### 2.2 Pre-existing Conditions and One-Year Missed Appointment Rates

#### 2.2.1 Data Source and Study Design

To examine the association between specific veteran-prevalent conditions and missed appointment rates, we identified Veterans who had at least two outpatient visits during calendar year 2023 (N = 5,545,723). Pre-existing conditions—including PTSD, TBI, Parkinson’s disease, and amputation—were defined using diagnosis codes recorded prior to January 1, 2023 (**Online Table 1**). Veterans without any of these conditions served as the reference group (**Online Figure 1B**).

**Table 1.** Missed Appointment Rates by Visit Modality.

| **Table 1. Missed Appointment Rates by Visit Modality** |  |  |  |  |  |
| --- | --- | --- | --- | --- | --- |
| **Modality** | **Canceled by**<br>**Clinic** | **Canceled by**<br>**Patients** | **Total Cancellation** | **No Show** | **Total**<br>**Missed** |
| VVC | 13% | 12% | 24% | 10% | 34% |
| Phone | 12% | 6% | 18% | 6% | 24% |
| F2F | 12% | 14% | 26% | 6% | 32% |
| CVT | 14% | 15% | 29% | 9% | 39% |
| Abbreviations: VVC = VA Video Connect (direct-to-home video visit); Phone = telephone visit (audio only); F2F = face-to-face (in-person) visit; CVT = Clinical Video Telehealth (video visit conducted at a VA or affiliated clinical site). |  |  |  |  |  |

#### 2.2.2 Statistical Analysis

We compared missed appointment rates across veterans with pre-existing condition groups. Adjusted associations between clinical conditions and missed appointment rates were estimated using regression models that controlled for age, sex, and race.

### 2.3 Missed Appointment Rates and Adverse Clinical Outcomes

#### 2.3.1 Data Source and Study Design

To evaluate the association between missed appointment rates and adverse clinical outcomes, we conducted two independent cohort analyses. We identified Veterans with a diagnosis of PTSD (N = 1,429,890) and, separately, Veterans with a diagnosis of TBI (N = 554,553) prior to 2023 who had at least two outpatient visits in 2023 and were alive as of December 31, 2023 (**Online Figure 1C**).

#### 2.3.2 Missed Appointment Measures

For patient-level analyses, missed appointment rates were calculated for each Veteran in 2023 as the number of missed appointments divided by the total number of scheduled appointments and were categorized into tertiles separately for each missed appointment type (no-show, clinic-initiated cancellation, and patient-initiated cancellation).

#### 2.3.2 Covariates

January 1, 2024, was used as the index date. Age was calculated as of the index date. Comorbidities for both the PTSD and TBI cohorts were identified using ICD codes recorded prior to the index date. To account for healthcare utilization, we also included each Veteran’s total outpatient visit count in 2023 as a separate covariate. Hospitalization status in the year preceding the index date was determined using inpatient data from the VHA and Medicare. Information on death and dates of death was obtained from the VA’s updated Death Ascertainment File.

#### 2.3.3 Clinical Outcomes

Primary outcomes included all-cause hospitalization, all-cause mortality, and a composite outcome of hospitalization or death within 1 year of January 1, 2024.

#### 2.3.4 Statistical Analysis

We estimated hazard ratios for the one year risks of hospitalization, death, and the composite outcome using time-to-event models, with tertiles of missed appointment rate as the primary exposures. Models adjusted for demographic characteristics, comorbidities, and prior hospitalization history.

## 3. Results

### 3.1 VA Missed Appointments Over Time and Across Facilities

Between January 1, 2000, and December 31, 2024, we identified 2.16 billion scheduled outpatient appointments with a valid status. Overall, 6.5% of appointments resulted in no-shows, and 25.4% were canceled, including both clinic-initiated and patient-initiated cancellations.

Substantial geographic and temporal variation in missed appointment rates was observed across VA facilities (**Figures 1 and 2**). Across facilities, no-show rates ranged from 3.5% to 14.1%, patient-initiated cancellation rates from 9.7% to 26.0%, and clinic-initiated cancellation rates from 8.5% to 17.9%.

**Figure 2.**
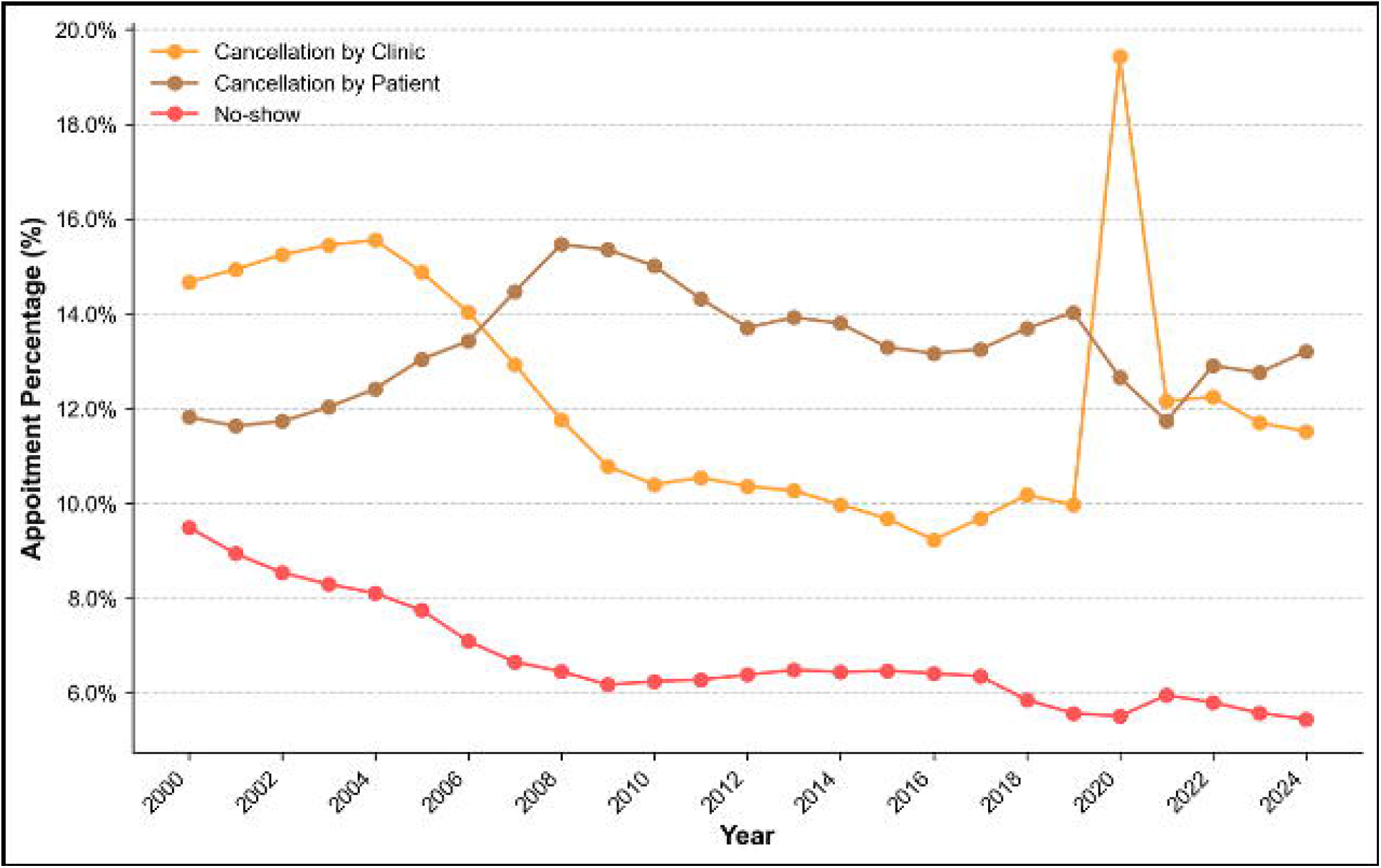
Percentage of no-show, clinic-initiated cancellation, and patient-initiated cancellation appointments in the VA, 2000–2024.

Temporal trends differed by appointment outcome. Clinic-initiated cancellation rates fluctuated over time and exhibited a pronounced peak in 2020, coinciding with the onset of the COVID-19 pandemic. In contrast, patient-initiated cancellation rates increased in the early 2000s, peaked around 2008–2009, and then gradually declined, stabilizing in the later study years. No-show rates declined steadily from 2000 to 2009, remained relatively stable through 2017, increased modestly during the pandemic, then declined again. The lowest observed no-show rate was 5.4% in 2024.

### 3.2 Missed Appointments by Visit Modality

Missed appointment rates varied meaningfully by visit modality. Telephone visits had the lowest overall missed appointment rate (24%), compared with face-to-face visits (32%), a difference driven primarily by substantially lower patient-initiated cancellation rates for telephone visits (6% vs 14%) (**Table 1**). Rates remained elevated for telehealth modalities requiring video, including VA Video Connect (34%) and Clinical Video Telehealth (39%).

### 3.3 Missed Appointments by Pre-existing Conditions

Among 5,545,723 Veterans with at least 2 outpatient visits in 2023, missed appointment rates varied by pre-existing clinical condition (**Table 2**). Veterans with Parkinson’s disease had the highest clinic-initiated cancellation rate (12.7%), whereas Veterans with TBI had the highest patient-initiated cancellation rate (13.5%). Veterans with TBI also exhibited the highest no-show rate (7.5%), which was approximately 1.6 times higher than the rate observed among Veterans without amputation, Parkinson’s disease, PTSD, or TBI.

**Table 2:**
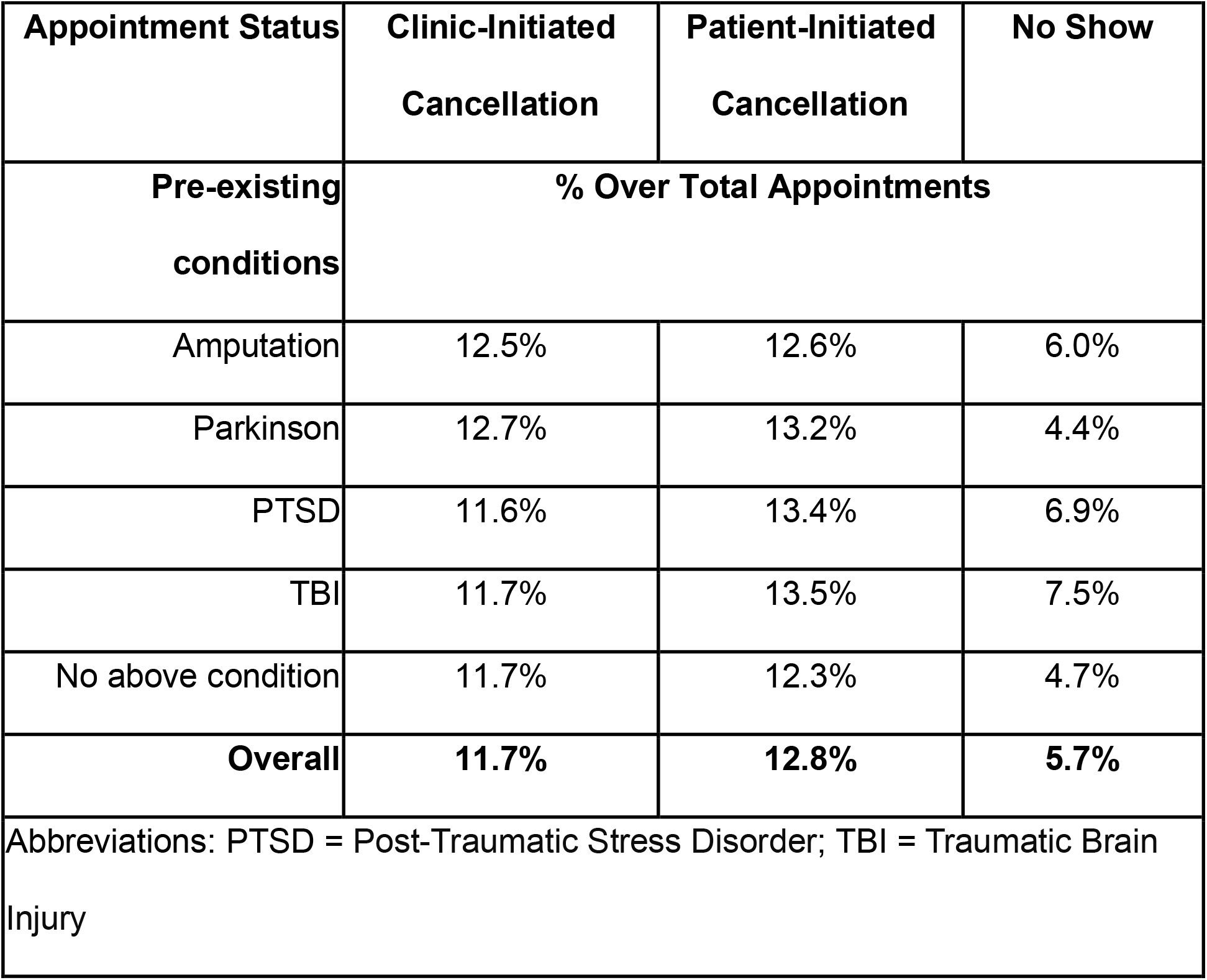
Missed appointment rates by pre-existing conditions and appointment status, 2023.

| **Appointment Status** | **Clinic-Initiated**<br>**Cancellation** | **Patient-Initiated**<br>**Cancellation** | **No Show** |
| --- | --- | --- | --- |
| **Pre-existing**<br>**conditions** | **% Over Total Appointments** |  |  |
| Amputation | 12.5% | 12.6% | 6.0% |
| Parkinson | 12.7% | 13.2% | 4.4% |
| PTSD | 11.6% | 13.4% | 6.9% |
| TBI | 11.7% | 13.5% | 7.5% |
| No above condition | 11.7% | 12.3% | 4.7% |
| **Overall** | **11.7%** | **12.8%** | **5.7%** |
| Abbreviations: PTSD = Post-Traumatic Stress Disorder; TBI = Traumatic Brain Injury |  |  |  |

After adjustment for age, sex, and race, pre-existing conditions, including PTSD, TBI, and Parkinson’s disease, remained significantly associated with higher missed appointment rates for both no-shows and cancellations compared with the reference group (all p < 0.05), indicating that the elevated rates among these clinical populations were not explained solely by demographic differences.

### 3.4 Missed Appointments and Clinical Outcomes

Baseline characteristics for 1,429,890 Veterans with PTSD and 554,553 Veterans with TBI are presented in **online Tables 2 and 3**. Overall, 8.4% of Veterans with PTSD experienced hospitalization or death within one year of follow-up; the corresponding proportion among Veterans with TBI was 10.9% (**online Table 4**)

**Table 3:** Hazard ratios for one-year risk of hospitalization or death across missed-appointment rate tertiles among (a) patients with PTSD and (b) patients with TBI.

| Outcomes | Death | Hospitalization | Death or Hospitalization |
| --- | --- | --- | --- |
|  | Hazard Ratio (95% CI) |  |  |
| a) Patients with PTSD |  |  |  |
| Clinic-Initiated Cancellation |  |  |  |
| Zero Cancellation | Reference |  |  |
| Cancellation Rate Tertile 1 | 0.79 (0.76-0.83) | 1.10 (1.08-1.13) | 1.00 (0.98-1.02) |
| Cancellation Rate Tertile 2 | 0.85 (0.82-0.89) | 1.10 (1.07-1.13) | 1.00 (0.98-1.02) |
| Cancellation Rate Tertile 3 | 1.01 (0.98-1.05) | 1.07 (1.04-1.09) | 1.03 (1.01-1.05) |
| Patient-Initiated Cancellation |  |  |  |
| Zero Cancellation | Reference |  |  |
| Cancellation Rate Tertile 1 | 0.89 (0.85-0.92) | 1.09 (1.07-1.12) | 1.02 (1.00-1.04) |
| Cancellation Rate Tertile 2 | 1.00 (0.96-1.03) | 1.04 (1.02-1.07) | 1.01 (0.99-1.03) |
| Cancellation Rate Tertile 3 | 1.30 (1.25-1.35) | 0.97 (0.95-1.00) | 1.00 (0.98-1.02) |
| No-show |  |  |  |
| Zero No-show | Reference |  |  |
| No-show Rate Tertile 1 | 1.10 (1.07-1.14) | 1.05 (1.03-1.07) | 1.01 (0.99-1.03) |
| No-show Rate Tertile 2 | 1.37 (1.32-1.42) | 1.01 (0.99-1.03) | 1.02 (1.00-1.04) |
| No-show Rate Tertile 3 | 1.91 (1.84-1.98) | 1.04 (1.02-1.06) | 1.07 (1.05-1.08) |
| b) Patients with TBI |  |  |  |
| Clinic-Initiated Cancellation |  |  |  |
| Zero Cancellation | Reference |  |  |
| Cancellation Rate Tertile 1 | 0.78 (0.74-0.82) | 1.43 (1.38-1.47) | 1.20 (1.17-1.23) |
| Cancellation Rate Tertile 2 | 0.86 (0.82-0.91) | 1.42 (1.37-1.46) | 1.21 (1.18-1.25) |
| Cancellation Rate Tertile 3 | 1.02 (0.97-1.06) | 1.25 (1.21-1.29) | 1.15 (1.12-1.18) |

| Patient-Initiated Cancellation |  |  |  |
| --- | --- | --- | --- |
| Zero Cancellation | Reference |  |  |
| Cancellation Rate Tertile 1 | **0.87 (0.83-0.92)** | **1.34 (1.30-1.38)** | **1.20 (1.17-1.23)** |
| Cancellation Rate Tertile 2 | 0.97 (0.92-1.01) | **1.23 (1.19-1.27)** | **1.14 (1.12-1.18)** |
| Cancellation Rate Tertile 3 | **1.24 (1.18-1.30)** | **1.11 (1.07-1.15)** | **1.12 (1.09-1.16)** |
| No-show |  |  |  |
| Zero No-show | Reference |  |  |
| No-show Rate Tertile 1 | **1.17 (1.12-1.22)** | **1.46 (1.43-1.50)** | **1.38 (1.35-1.41)** |
| No-show Rate Tertile 2 | **1.30 (1.24-1.37)** | **1.41 (1.37-1.45)** | **1.39 (1.36-1.42)** |
| No-show Rate Tertile 3 | **1.62 (1.55-1.70)** | **1.63 (1.58-1.67)** | **1.65 (1.61-1.69)** |
| *Hazard ratios indicating a statistically significant increased risk are bolded in red; those indicating a statistically significant decreased risk are bolded in green. |  |  |  |

When Veterans were categorized into tertiles based on their 2023 no-show rate (low, medium, high), higher no-show tertiles were consistently associated with increased risk of hospitalization, death, and the composite outcome during the subsequent year (**Tables 3a and 3b**).

Among Veterans with PTSD, compared with Veterans with zero no-shows (reference group), adjusted mortality risk increased stepwise across no-show tertiles among those with ≥1 no-show, with hazard ratios of 1.10 (95% CI 1.07–1.14) for the lowest tertile and 1.91 (95% CI 1.84–1.98) for the highest tertile. Associations for clinic-initiated and patient-initiated cancellations were more heterogeneous than those for no-shows. However, higher cancellation tertiles generally corresponded to elevated risks, with the most consistent risk increases observed among Veterans with TBI.

## 4. Discussion

### 4.1 Significance

Unattended appointments, including both no-shows and cancellations, may disrupt the continuity of care,^10^ strain clinical resources, and contribute to substantial financial losses.^11,12^ This study offers a comprehensive evaluation of missed appointment patterns among Veterans receiving care within the VA healthcare system, aiming to better inform efforts to improve access and care coordination.

Missed visits may also contribute to the worsening of existing health conditions by delaying essential treatments, monitoring, and supportive services.^13^ Our findings indicate that higher levels of missed appointments are associated with poorer outcomes among Veterans with pre-existing conditions such as PTSD and TBI, underscoring the need for targeted strategies to maintain engagement in care among these high-risk groups.

### 4.2 Temporal Variations in Missed Appointments

We observed changes in missed appointment patterns over time that likely reflect both patient behavior and system-level factors. Since the mid-2010s, VA has implemented multiple initiatives to reduce missed appointments and improve access, such as the National Initiative to Reduce Missed Opportunities (NIRMO) and the standardization of recall reminder systems,^14-16^ which may partly explain the decline in no-show rates over time.

During the COVID-19 pandemic, the rapid expansion of telehealth^17-19^ may have enabled some veterans to attend appointments who would otherwise have faced barriers to traveling to clinics. At the same time, disruptions in routine care, including clinic closures, staffing shortages, and patient anxiety, may have led to increased missed visits by others. After the pandemic, the continued use of telehealth, along with system-wide efforts to improve access, may also have contributed to a partial recovery. Our data comparing the missed-appointment rate in visit modalities also suggests that flexible care-delivery models, such as telephone visits, may be associated with improved appointment adherence, while technology-dependent video modalities may continue to face persistent barriers to completion.

### 4.3 Geographic and Facility-Level Variation in Missed Appointments

Our findings also showed that missed appointment rates varied across VA facilities and regions. This likely reflects differences in patient populations as well as local system factors, including travel distance, transportation barriers, and the availability of specialty clinicians or clinics. In addition, heterogeneity in visit modality (in-person, telephone, or video) and local telehealth resources may contribute to observed differences across facilities and regions. ^20^

What is more, the type of care matters. Missed appointments may differ substantially between primary care and specialty clinics, such as neurology or mental health settings.^20^ These settings have distinct scheduling practices, visit purposes, and patient needs, which may influence cancellation and no-show patterns. For example, primary care within the VHA is often supported by enterprise standards and more consistent clinic structures, whereas specialty care may involve greater variability in clinic templates, staffing, and availability. Understanding how local clinic structure, staffing, and modality mix interact with patient barriers will be important for designing solutions tailored to the regional context.

### 4.4 Missed Appointments and Pre-Existing Conditions

Our analysis also showed that patients with high-need pre-existing conditions such as TBI, PTSD, Parkinson’s disease, or amputation were more likely to have missed appointments, potentially reflecting greater clinical complexity, functional limitations, or barriers to care. Patients with pre-existing conditions may also have higher healthcare utilization and, therefore, more scheduled visits, which could increase the likelihood of missed appointments. To account for this, we adjusted for healthcare utilization by including each Veteran’s total outpatient visit count in 2023 as a covariate in our models. However, differences in utilization alone cannot fully explain the variation observed across diagnostic groups. Notably, we observed distinct patterns based on the specific condition: Veterans with TBI exhibited the highest no-show rates, whereas those with Parkinson’s disease experienced the highest cancellation rates. These discrepancies highlight the unique obstacles faced by different patient populations, such as the cognitive or memory impairments often associated with TBI^21^ versus the physical mobility inherent in Parkinson’s care.

### 4.5 Missed Appointments and Adverse Outcomes

This study is among the first to examine the association between missed appointments and adverse outcomes among Veterans with pre-existing conditions (specifically PTSD and TBI). By examining graded levels of both no-shows and cancellations, our results show that a higher proportion of missed visits is associated with increased risk of adverse outcomes among patients with these conditions. The association was most consistent for no-shows, supporting their potential role as a clinically meaningful risk indicator rather than isolated events. Repeated no-shows may signal difficulty maintaining engagement with care over time.^22^ Because Veterans with worsening health may miss appointments shortly before hospitalization, reverse causation is possible. Even if missed visits are not the direct cause of worse outcomes, they appear to serve as a warning sign that a veteran may be at higher clinical risk and in need of additional support.^23,24^

In contrast, associations for cancellations were more heterogeneous than those for no-shows, particularly for mortality. This pattern may reflect, in part, clinical triage and scheduling dynamics—for example, lower-acuity follow-up visits may be more likely to be canceled (by clinics or patients), whereas higher-risk patients may be rescheduled more promptly or managed through other channels (e.g., urgent care or inpatient care). Nevertheless, it is important to note that within the VA, cancellations and no-shows are sometimes grouped together as “missed opportunities,” but our findings suggest that combining these behaviors may obscure meaningful differences in their clinical implications.

### 4.6 Implications

These observations underscore the potential need for condition-specific, as well as patient-tailored, strategies to reduce missed appointments. While modest cancellation rates may reflect adaptive scheduling and pose limited harm, persistently high no-show rates and late clinic cancellations represent lost therapeutic opportunities and should be prioritized in intervention design. Tailored outreach—potentially combining reminder systems, telehealth flexibility, and social support—could help reduce disparities in access and improve downstream health outcomes.

Missed appointments may also allow otherwise manageable health issues to worsen. Diagnostic testing may be delayed, medications may not be initiated or adjusted, and routine follow-up becomes more difficult to maintain. Over time, care can become fragmented. For Veterans living with chronic conditions—such as cognitive impairment, mobility limitations, or mental health disorders—transportation barriers and other functional challenges may render standard scheduling models less effective. More flexible, condition-tailored approaches may help keep these Veterans engaged in care.^25^

### 4.7 Limitations

Several limitations warrant acknowledgment. First, the analysis did not incorporate the interval between appointment scheduling and cancellation, nor did it distinguish among primary care, specialty, and mental health visits. We also could not differentiate cancellations that were subsequently rebooked from those that were not, which may have different clinical implications.

Second, our classification of no-shows and cancellations relied on administrative appointment status fields. A substantial number of appointments were marked as “no action taken” or had unclear status (N = 103,461,999; 4.5% of all appointments) and were excluded from the analytic dataset, which may introduce misclassification or selection bias if these statuses are associated with patient risk or facility practices. In addition, we assumed appointments were completed when no appointment status was recorded; this assumption may not always be correct and could bias estimates toward undercounting missed visits.

Third, data on key socioeconomic factors—such as travel distance and transportation barriers—were not available. Additionally, while the VA population increasingly includes younger and female Veterans, it remains predominantly composed of older men, which may limit generalizability.

Fourth, although models adjusted for total outpatient visit count, residual confounding by overall utilization burden may remain; planned work will examine stratified analyses among high- and low-utilizing Veterans. Finally, as with all observational studies, causal relationships cannot be established. Missed appointments may reflect underlying illness severity, social vulnerability, or both.

### 4.8 Future Work

We plan to expand the diagnostic and temporal risk-factor profile by integrating geographic, social, and behavioral variables within the VA dataset. Advanced modeling approaches, including causal inference techniques, will be used to better distinguish predictors from downstream consequences. In addition, machine-learning methods will be applied to identify high-risk individuals early, enabling proactive scheduling adjustments and reducing avoidable lapses in care.^**26**,**27**^

The primary goal is to detect Veterans who may be struggling to engage with care at an early stage and respond in near real time. By tailoring scheduling practices and visit options to individual needs, the VA may reduce missed appointments and improve continuity of care. Preventing care from falling through the cracks can improve long-term health outcomes.

## Data Availability

The data analyzed in this study were obtained through the US Department of Veterans Affairs (VA) Corporate Data Warehouse (CDW). These data are accessed through the Veterans Affairs Informatics and Computing Infrastructure (VINCI), a secure computing environment located behind the VA firewall. Access to CDW data is limited to authorized users and requires specific approvals, with all use governed by VA policies and regulations related to data security, privacy, and responsible data stewardship.

## Statement of AI Assistance

This manuscript was reviewed for grammar, syntax, and language clarity using ChatGPT (OpenAI, GPT-5.4 Thinking). The tool was used solely to support language refinement. All scientific content, interpretation, and conclusions were developed and verified by the authors.

## Ethical Approval

The study was approved by the Institutional Review Board of the Washington DC Veterans Affairs Medical Center.

