## Supplementary material for "Missed Appointments and Associations with Clinical Outcomes in A Large National Healthcare System": online supplements

**Online Figure 1. Flowchart of study cohort assembly**

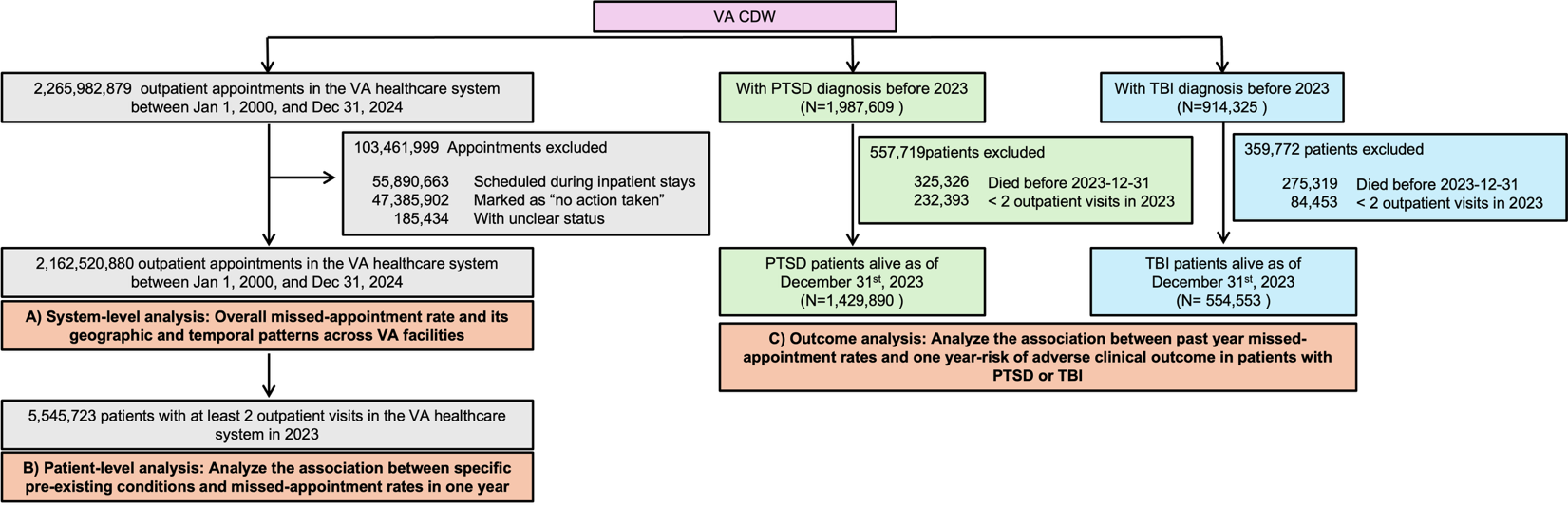

| **Online Table 1: International Classification of Diseases (ICD) codes** | | |
| --- | --- | --- |
| **Condition** | **ICD-9CM** | **ICD-10CM** |
| Amputation | 885.x, 886.x, 887.x, 895.x, 896.x, 897.x, V49.6, V49.7, 997.60 | S08.x, S48.x, S58.x, S68.x, S78.x, S88.x, S98.x, T87.x, Z89.x, S28.1x, S28.2x, S38.1x, |
| Parkinson | 332.x, 331.82 | G20.x, G31.83 |
| PTSD | 309.81x | F43.1x |
| TBI | 310.2, 800.x, 801.x, 803.x, 804.x, 850.x, 851.x, 852.x, 853.x, 854.x, 905.0, 907.0, 950.1, 950.2, 950.3, 959.01, 959.9, V15.52 | S02.0x, S02.1x, S02.8x, S02.9x, S04.02x, S04.03x, S04.04x, S06.x, S07.1x, Z87.82 |

| **Online Table 2: Baseline Characteristics of Veterans With PTSD** | |
| --- | --- |
| **Characteristic** | **No. (%) or Mean (SD)** |
| **Overall** | 1,429,890 |
| **Missed Appointment Rate Groups (2023)** | |
| **Clinic-initiated cancellation rate** | |
| No Clinic-initiated cancellation (0%) | 360,317 (25.2) |
| Tertile 1 (>0%–10%) | 373,087 (26.1) |
| Tertile 2 (>10%–16%) | 332,846 (23.3) |
| Tertile 3 (>16%) | 363,094 (25.4) |
| **Patient-initiated cancellation rate** | |
| No patient-initiated cancellation (0%) | 359,601 (25.1) |
| Tertile 1 (>0%–10%) | 316,181 (22.1) |
| Tertile 2 (>10%–20%) | 397,577 (27.8) |
| Tertile 3 (>20%) | 355,985 (24.9) |
| **No-show rate** |  |
| No no-show (0%) | 682,687 (47.7) |
| Tertile 1 (>0%–7%) | 255,443 (17.9) |
| Tertile 2 (>7%–15%) | 234,745 (16.4) |
| Tertile 3 (>15%) | 256,469 (17.9) |
| **Age, years** | 55.1 (15.6) |
| **Sex** |  |
| Female | 222,861 (15.6) |
| Male | 1,207,029 (84.4) |
| **Race** |  |
| White | 905,099 (63.3) |
| Black | 334,573 (23.4) |
| Asian | 21,187 (1.5) |
| American Indian or Alaska Native | 15,436 (1.1) |
| Native Hawaiian or Other Pacific Islander | 16,335 (1.1) |
| Other or Unknown | 137,260 (9.6) |
| **Ethnicity** |  |
| Hispanic | 142,744 (10.0) |
| Non-Hispanic | 1,209,648 (84.6) |
| Other or Unknown | 77,498 (5.4) |
| **Substance Use Disorders** |  |
| Alcohol use disorder | 520,156 (36.4) |
| Opioid use disorder | 95,667 (6.7) |
| Tobacco use disorder | 636,969 (44.5) |
| **Mental Health Conditions** |  |
| Anger-related disorders | 359,260 (25.1) |
| Anxiety disorders | 976,070 (68.3) |
| Attention-deficit/hyperactivity disorder | 104,914 (7.3) |
| Bipolar disorder | 171,963 (12.0) |
| Disruptive behavior disorders | 36,480 (2.6) |
| Major depressive disorder | 1,058,359 (74.0) |
| Minor depression | 742,302 (51.9) |
| Psychosis (other) | 88,917 (6.2) |
| Schizophrenia | 42,017 (2.9) |
| Sleep disorders | 1,047,781 (73.3) |
| Suicidal ideation | 216,855 (15.2) |
| Suicide attempt | 53,800 (3.8) |
| **Medical Comorbidities** |  |
| Cancer | 173,592 (12.1) |
| Cardiac Procedure | 105,746 (7.4) |
| Chronic pulmonary disease | 438,364 (30.7) |
| Dementia | 48,630 (3.4) |
| Dementia | 48,630 (3.4) |
| Heart failure | 125,249 (8.8) |
| Hyperlipidemia | 890,721 (62.3) |
| Hypertension | 845,100 (59.1) |
| Myocardial infarction | 89,290 (6.2) |
| Peripheral vascular disease | 163,357 (11.4) |
| Renal disease | 182,679 (12.8) |
| **Hospitalization in 2023** | 106,922 (7.5) |

| **Online Table 3. Baseline Characteristics of Veterans With TBI** | |
| --- | --- |
| **Characteristic** | **No. (%) or Mean (SD)** |
| **Overall** | 554,553 |
| **Missed Appointment Rate Groups** |  |
| **Clinic-initiated cancellation rate** |  |
| No Clinic-initiated cancellation (0%) | 154,941 (27.9) |
| Tertile 1 (>0%–10%) | 150,809 (27.2) |
| Tertile 2 (>10%–16%) | 115,429 (20.8) |
| Tertile 3 (>16%) | 133,374 (24.1) |
| **Patient-initiated cancellation rate** |  |
| No patient-initiated cancellation (0%) | 152,978 (27.6) |
| Tertile 1 (>0%–10%) | 126,988 (22.9) |
| Tertile 2 (>10%–20%) | 153,399 (27.7) |
| Tertile 3 (>20%) | 121,188 (21.9) |
| **No-show rate** |  |
| No no-show (0%) | 262,158 (47.3) |
| Tertile 1 (>0%–7%) | 97,005 (17.5) |
| Tertile 2 (>7%–16%) | 97,695 (17.6) |
| Tertile 3 (>16%) | 97,695 (17.6) |
| **Age, years** | 54.8 (15.8) |
| **Sex** |  |
| Female | 55,563 (10.0) |
| Male | 498,990 (90.0) |
| **Race** |  |
| White | 388,795 (70.1) |
| Black | 95,495 (17.2) |
| Asian | 7,640 (1.4) |
| Native Hawaiian or Other Pacific Islander | 6,319 (1.1) |
| American Indian or Alaska Native | 6,464 (1.2) |
| Other or Unknown | 49,840 (9.0) |
| **Ethnicity** |  |
| Non-Hispanic | 471,196 (85.0) |
| Hispanic | 56,276 (10.1) |
| Other or Unknown | 27,081 (4.9) |
| **Marital Status** |  |
| Married | 271,001 (48.9) |
| Single or Never Married | 103,945 (18.7) |
| Widowed, Divorced, or Separated | 172,257 (31.1) |
| Unknown | 7,350 (1.3) |
| **Substance Use Disorders** |  |
| Alcohol use disorder | 261,152 (47.1) |
| Opioid use disorder | 158,573 (28.6) |
| Tobacco use disorder | 248,899 (44.9) |
| **Mental Health Conditions** |  |
| Anxiety disorders | 341,116 (61.5) |
| Attention-deficit/hyperactivity disorder | 43,132 (7.8) |
| Bipolar disorder | 74,208 (13.4) |
| Major depressive disorder | 354,933 (64.0) |
| Minor depression | 290,791 (52.4) |
| Other psychotic disorders | 28,650 (5.2) |
| Panic disorders | 51,102 (9.2) |
| Post-traumatic stress disorder | 352,431 (63.6) |
| Schizophrenia | 27,264 (4.9) |
| Sleep disorders | 396,010 (71.4) |
| Suicidal ideation | 89,352 (16.1) |
| **Neurologic Conditions** |  |
| Narcolepsy | 3,574 (0.6) |
| Other neurologic disorders | 444,163 (80.1) |
| Parkinson disease | 8,749 (1.6) |
| **Hospitalization in 2023** | 49,037 (8.8) |

| **Online Table 4. One-Year Clinical Outcomes in 2024 Among Veterans With PTSD and TBI** | | |
| --- | --- | --- |
| **Outcome** | **PTSD (N = 1,429,890)** | **TBI (N = 554,553)** |
| **Hospitalization** | 98,632 (6.9) | 48,773 (8.8) |
| **Death** | 27,250 (1.9) | 15,226 (2.7) |
| **Death or Hospitalization** | 119,627 (8.4) | 60,187 (10.9) |
| Abbreviations: PTSD = Post-Traumatic Stress Disorder; TBI = Traumatic Brain Injury | | |
